# Trends in the 30-year span of Noninfectious Cardiovascular Implantable Electronic Device Complications in Olmsted County

**DOI:** 10.1101/2023.05.09.23289751

**Authors:** Gurukripa N. Kowlgi, Vaibhav Vaidya, Ming-Yan Dai, Rahul Mishra, David O. Hodge, Abhishek J. Deshmukh, Siva K. Mulpuru, Paul A. Friedman, Yong- Mei Cha

## Abstract

**Background:** Cardiovascular implantable electronic devices (CIEDs) such as permanent pacemakers, implantable cardioverter-defibrillators, and cardiac resynchronization therapy devices alleviate morbidity and mortality in various diseases. There is a paucity of real-world data on CIED complications and trends.

**Objectives:** Describe trends in noninfectious CIED complications over the past three decades in Olmsted County.

**Methods:** The Rochester Epidemiology Project is a medical records linkage system comprising records of over 500,000 residents of Olmsted County from 1966-current. CIED implants between 1988-2018 were determined. Trends in noninfectious complications within 30 days of implant were analyzed.

**Results:** 175 out of 2536 (6.9%) patients who received CIED experienced device complications. 3.8% of the implants had major complications requiring intervention. Lead dislodgement was the most common (2.9%), followed by hematoma (2.1%). Complications went up from 1988 to 2005, then showed a downtrend until 2018, driven by a decline in hematomas in the last decade (p<0.01). Those with complications were more likely to have prosthetic valves. Obesity appeared to have a protective effect in a multivariate regression model. The mean Charlson comorbidity score has trended up over the 30 years.

**Conclusions:** Our study describes a real-world trend of CIED complications over three decades. Lead dislodgements and hematomas were the most common complications. Complications have declined over the last decade due to safer practices and a better understanding of anticoagulant management.

## Introduction

Cardiac implantable electronic devices (CIEDs) such as the permanent pacemaker (PPM), implantable cardioverter-defibrillator (ICD), and cardiac resynchronization therapy (CRT) systems are ubiquitously employed for managing a variety of cardiac arrhythmia and comorbidities. In recent years, we have witnessed an expansion in the indications for CIED implantation^1–4^. CIEDs have augmented patient quality of life, reduced hospitalization length, and reduced mortality^5–7^.

Notwithstanding substantial technological improvements, complications involving CIED implants remain a significant issue^8–10^. Acute complications of CIED implants include lead dislodgement, perforation, pneumothorax, pocket hematoma, infection, and upper extremity deep vein thrombosis (DVT)^8–11^. With an increase in life expectancy and expanding CIED implant indications, implant-related adverse events are anticipated^12–14^. Device complications are critical to detect and manage to improve patient safety, prevent reoperations, reduce the length of hospital stay, and mitigate downstream healthcare costs^15, 16^.

The majority of what we know about CIED complications is curated from secondary analysis of randomized trials^10, 17^, registry-based studies^18, 19^, or hospital-based databases^15, 20^. Administrative-claims-based databases have been shown to have low specificity in determining true CIED complications ^21^. There exists a paucity of real-life population-based data on the incidence, trends, and predictors of CIED complications. There exists a paucity of real-life population-based data on the incidence, trends, and predictors of CIED complications. Our group has previously published data on the trends of CIED infections ^14^. In the present study, we present retrospective population-based data on the noninfectious mechanical CIED complications between 1988 and 2018.

## Methods

### Data source

Our study was approved by the Mayo Clinic Institutional Review Board and received proper ethical oversight. Data were obtained from the Rochester Epidemiology Project (REP). The REP is a medical records linkage system containing medical records of all residents of Olmsted County, MN, from January 1, 1966, to the present, consisting of follow-up data on more than 500,000 unique individuals. Patient demographics, diagnostic codes such as the international classification of diseases 9^th^ and 10^th^ revision (ICD-9/10-CM) codes, and surgical procedure codes are recorded for all individuals. Paper and electronic medical records of these individuals are available for the generation of additional data. The REP permits the collection of population-based data and has been used to define the incidence of various medical conditions. It consists of two major hospital systems in Olmsted County (Mayo Clinic and Olmsted Medical Center) and smaller practices.

### Study population

The REP database was used to determine all patients who received CIED implantation between 1988 and 2018. During the study period, Mayo Clinic and Olmsted Medical Center were the only two institutions in Olmsted County performing CIED implantation and follow-up. There were four device implanters in the early 1990s and ten in 2018. Patients receiving CIEDs were determined based on ICD-9 CM and ICD-10 PCS codes. Data on subsequent device upgrades from PPM to ICD or CRT or ICD to CRT were collected using ICD-9 and 10 codes and manually verified. For this study, we only included procedures that involved intracardiac lead implantation. Thus, subcutaneous ICDs, generator change, and device extraction without lead implantation/revision were excluded. Data regarding baseline demographics and comorbidities were obtained from the REP and abstracted from the medical records. The patients’ charts with device complications were manually reviewed to confirm the complications. The Charlson comorbidity index is a score derived from multiple risk factors that predict 1-year survival and can be used as a surrogate for comorbidity. CCI was calculated for all patients at the time of the CIED implant. The unique complications were defined by a composite of ICD-9 and ICD-10 diagnosis codes, summarized in the **Supplemental Tables 1A-E**. For lead dislodgement, we used codes for a lead revision within 30 days and confirmed lead dislodgement through manual chart review. The clinical definitions for post-implant complications for this study are as follows. Notably, repeat surgical intervention was not a criterion to code the complications, and even those that were conservatively managed were included.

<u>Perforation:</u> Any new pericardial effusion, with or without hemodynamic compromise.

<u>Hematomas:</u> Any pocket bleeding that led to interruption of oral anticoagulant therapy, prolonged hospitalization > 24 hours.

<u>Pneumothorax:</u> Any pneumothorax detected on post-procedure chest radiography

<u>Lead dislodgement:</u> Included macro-dislodgement as evident on a chest radiograph, and micro-dislodgement that were detected based on acute rise in capture thresholds.

### Statistical analysis

Normally distributed continuous variables and expressed as mean ± standard deviation. The overall frequency of CIED complications was determined using the number of complications divided by the population at risk (device implants) in Olmsted County in the same period. Trends in complications over time, across age groups, and between genders are estimated using Poisson regression models. Comparisons between implant groups for categorical factors were completed using Chi-square tests. Continuous factors were compared between groups using the analysis of Variance. Overall survival was estimated using the Kaplan-Meier method. These curves were compared between groups using log-rank tests. P-values less than 0.05 were considered significant. All analysis was completed using SAS version 9.4.

## Results

### Population characteristics

Between 1988 and 2018, there were 2536 CIED initial implants in Olmsted County, including 1927 PPMs (single and dual-chamber), 440 ICDs, and 169 CRT (pacemaker and defibrillator) devices. The mean age at device implantation was 73.9 ± 14.3 years. Women comprised 42.1% of all CIED recipients in the study. 197 complications were seen in 175 (6.9%) patients. There was a significant difference in some baseline characteristics among patients with and without CIED complications (**Table 1**). Patients with CIED complications were more likely to have prosthetic valves and less likely to be obese or diabetic. Of the 197 complications, there were 75 (2.9%) lead dislodgements, 53 (2.1%) pocket hematomas, 42 (1.6%) pneumothoraces, and 27 (1.1%) cardiac perforations (**Table 2**). Patients with cardiac perforation were more likely to be female and less likely to have cardiomyopathy (**Table 3**). There was a higher frequency of cardiomyopathy, prosthetic valves, and stroke in those patients who suffered pocket hematomas. Patients with pneumothoraces were older, more likely to be females, and less likely to have obesity or cardiomyopathy. There was no demographic predisposition to lead dislodgement.

**Table 1:**
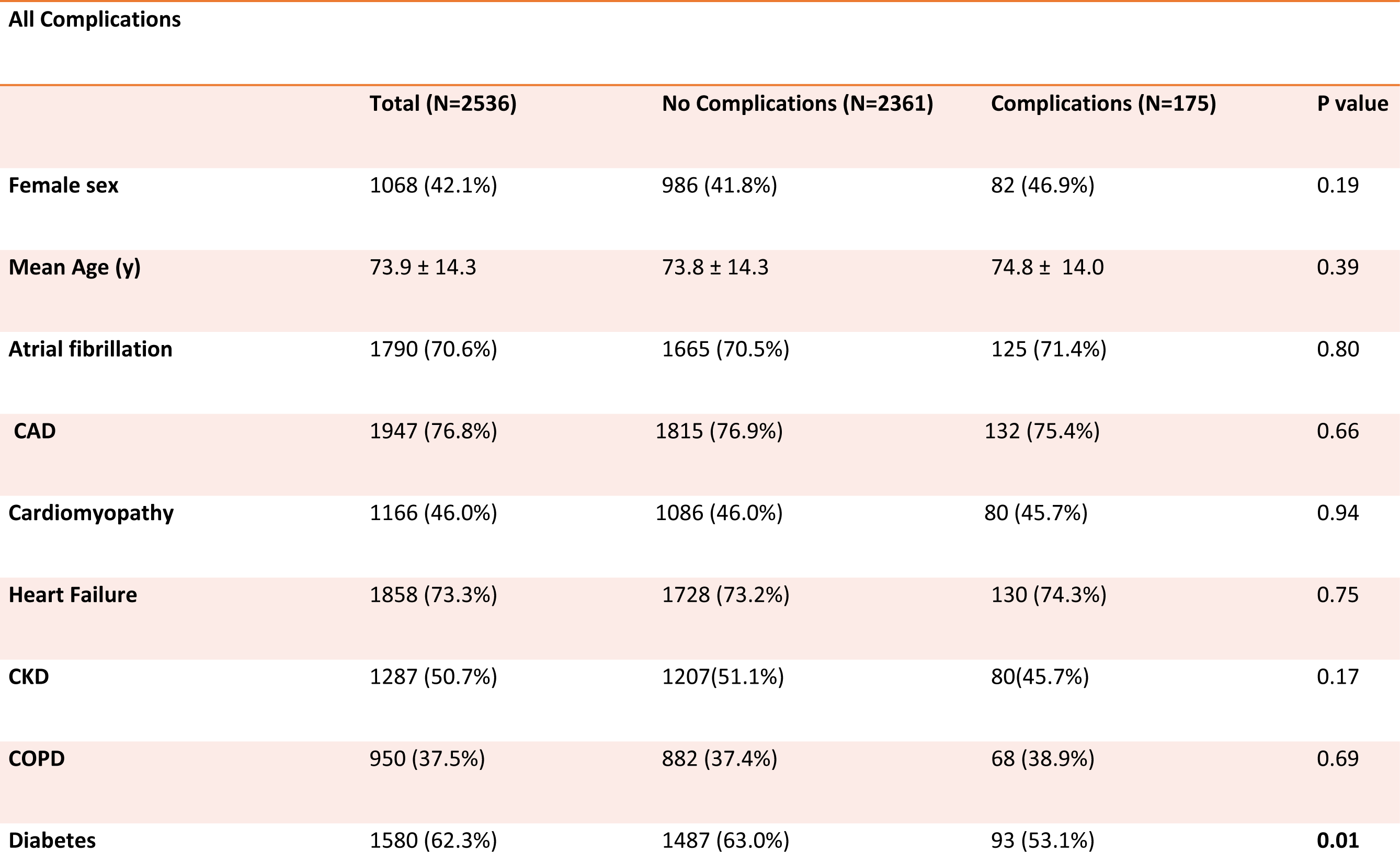
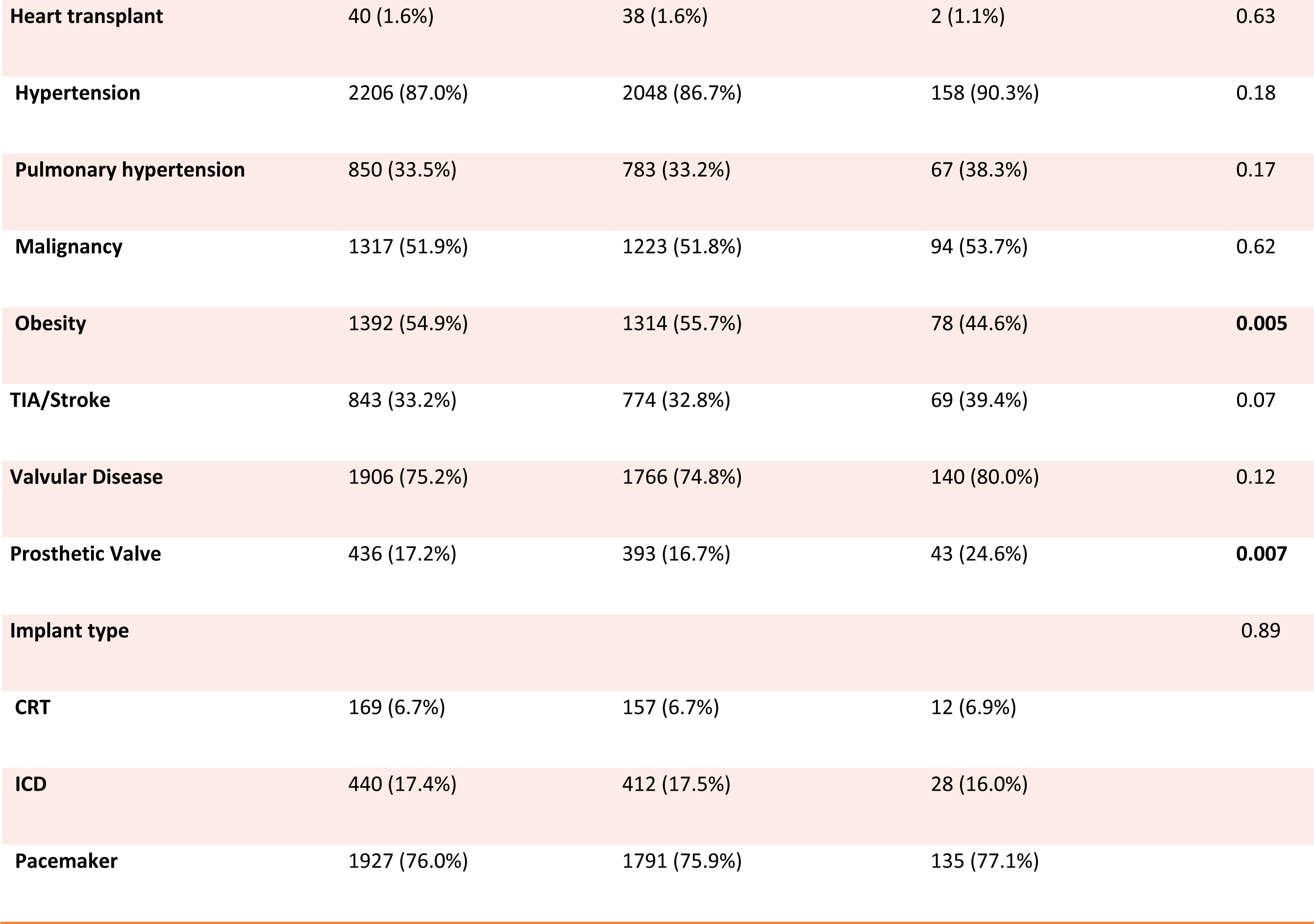
Baseline demographics for all patients.

**Table 2:**
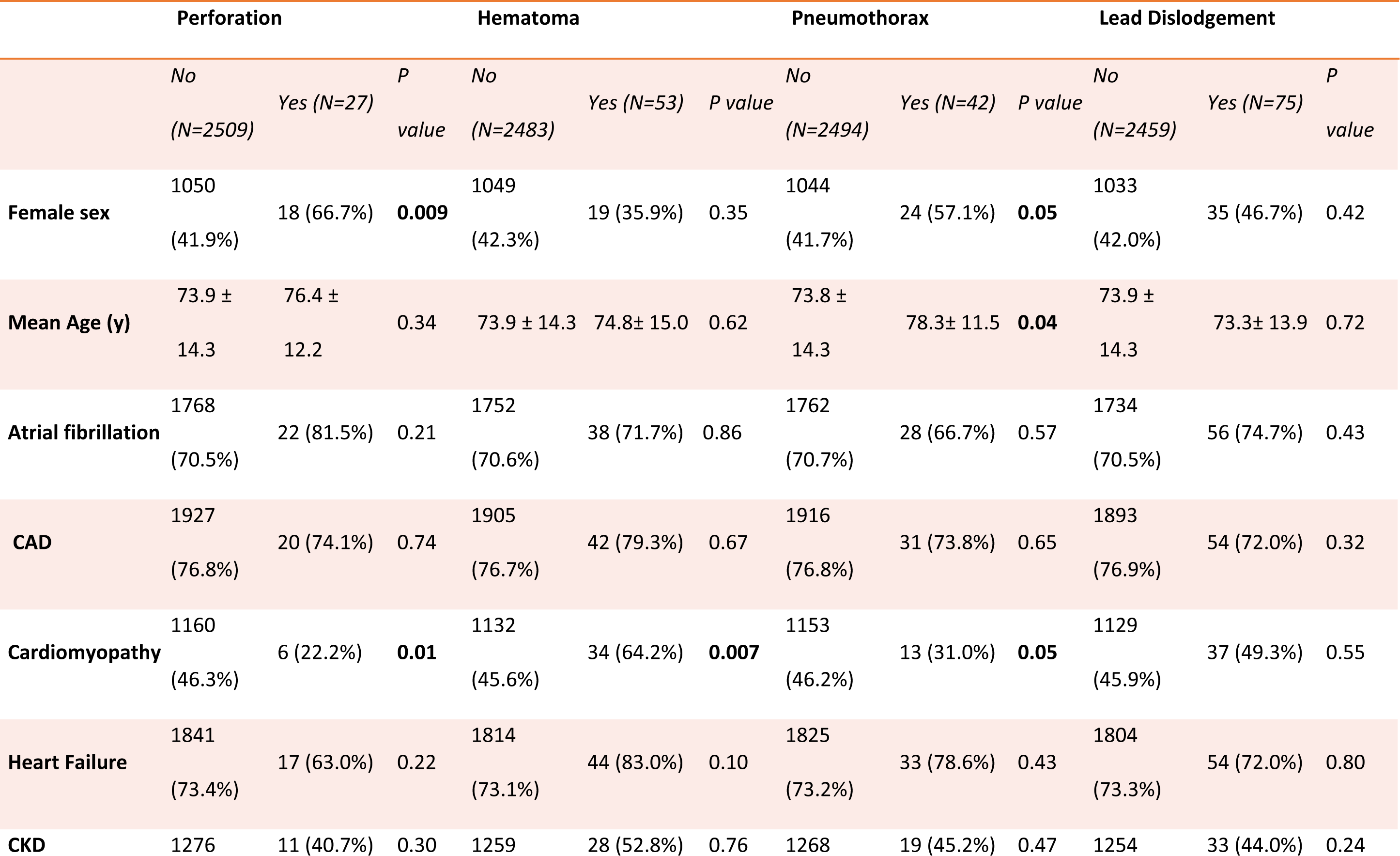
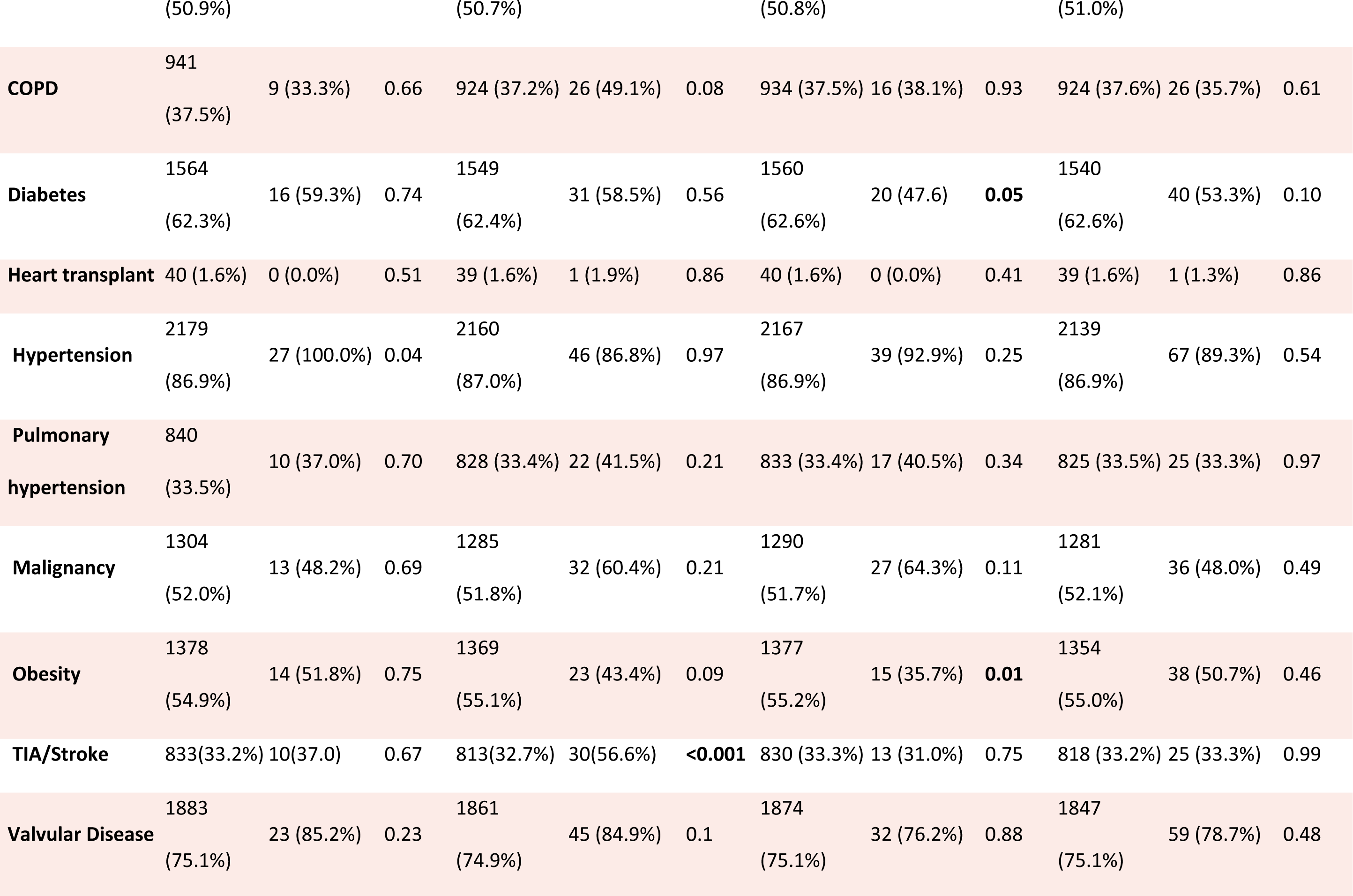
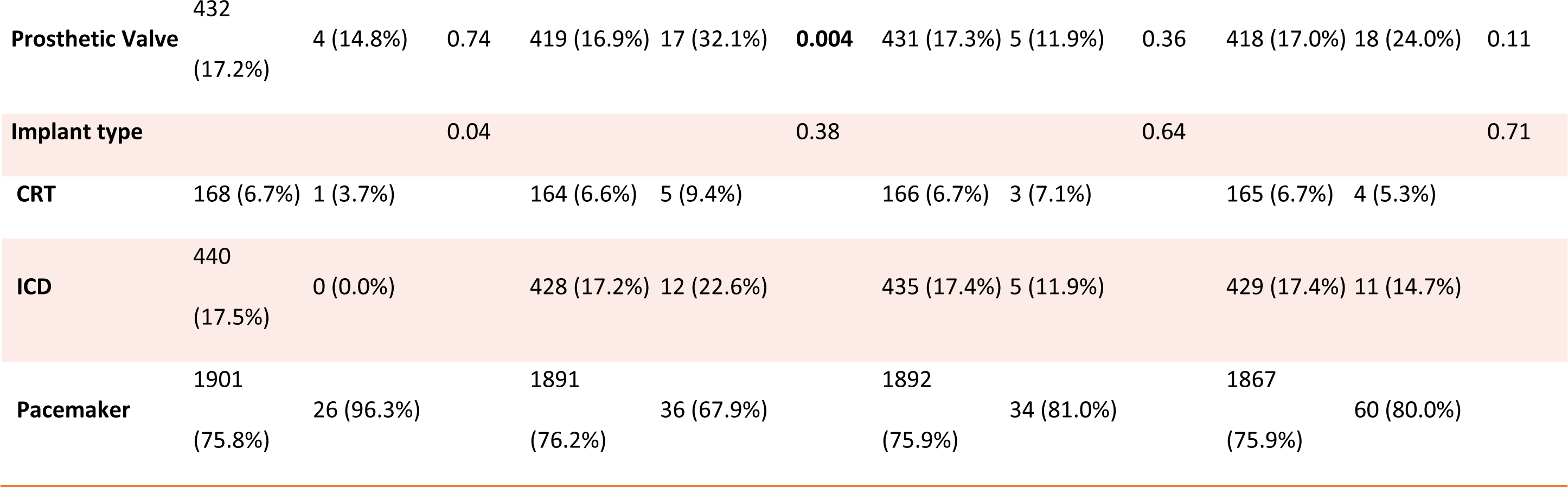
Baseline demographics by type of complication.

**Table 3:** Distribution of complications:

| Complications | Number (% of total implants) |
| --- | --- |
| Lead dislodgement | 75 (2.9%) |
| Hematoma | 53 (2.1%) |
| Pneumothorax | 42 (1.6%) |
| Perforation | 27 (1.1%) |

### Summary of complications

Of the patients with complications, 97 (3.8% of all implants) were classified as major complications requiring a procedural intervention as described. The number of complications per patient was one (133/175), two (18/175), or three (2/175). Over three-quarters of the patients with complications (135/175) had PPM implants. Sixteen percent of the patients with complications (28/175) underwent ICD implantation, and the remaining 6.9% (12/175) occurred in the CRT group. The complication rates in pacemakers, ICD, and CRT were 7%, 6.4%, and 7.1% (p=NS).

Of the 27 (1.1%) patients who had perforations, about half required pericardiocentesis (13/27). Only 5.7% (3/53) of the patients who developed hematomas required pocket re-entry to resolve the bleeding. The majority (88.7%) of the patients who developed hematomas were on either anticoagulation or antiplatelet medications or both, with 16.9% (9/53) receiving periprocedural heparin (all before 2013). More than two-thirds [69.1%, (29/42)] of patients with pneumothoraces were managed conservatively, and the rest required chest tube placement temporarily before resolution. Of the 75 patients with lead dislodgement, 35 (46.7%) had atrial lead dislodgement, 35 (46.7%) had their right ventricular lead dislodged, and 5 (5.3%) had their coronary sinus lead dislodged. Ninety-two percent of patients (69/75) had to have their dislodged lead revised.

### Temporal trends in CIED complications

Temporal trends were compared between five intervals: 1988-93, 94-99, 2000-2005, 2006-11, and 2012-18. There was a significant change in the proportion of complications over time (**Figure 1**). The total complications increased between 1988-93 to 2000-05; from 2.1% to 10.1%; and subsequently decreased to 7.7% in 2012-2018, p=0.001. Similar trends were noted for hematomas, p<0.01. There were variations in the frequencies of patients with perforations, pneumothoraces, and lead dislodgement; however, these were not statistically significant.

**Figure 1:**
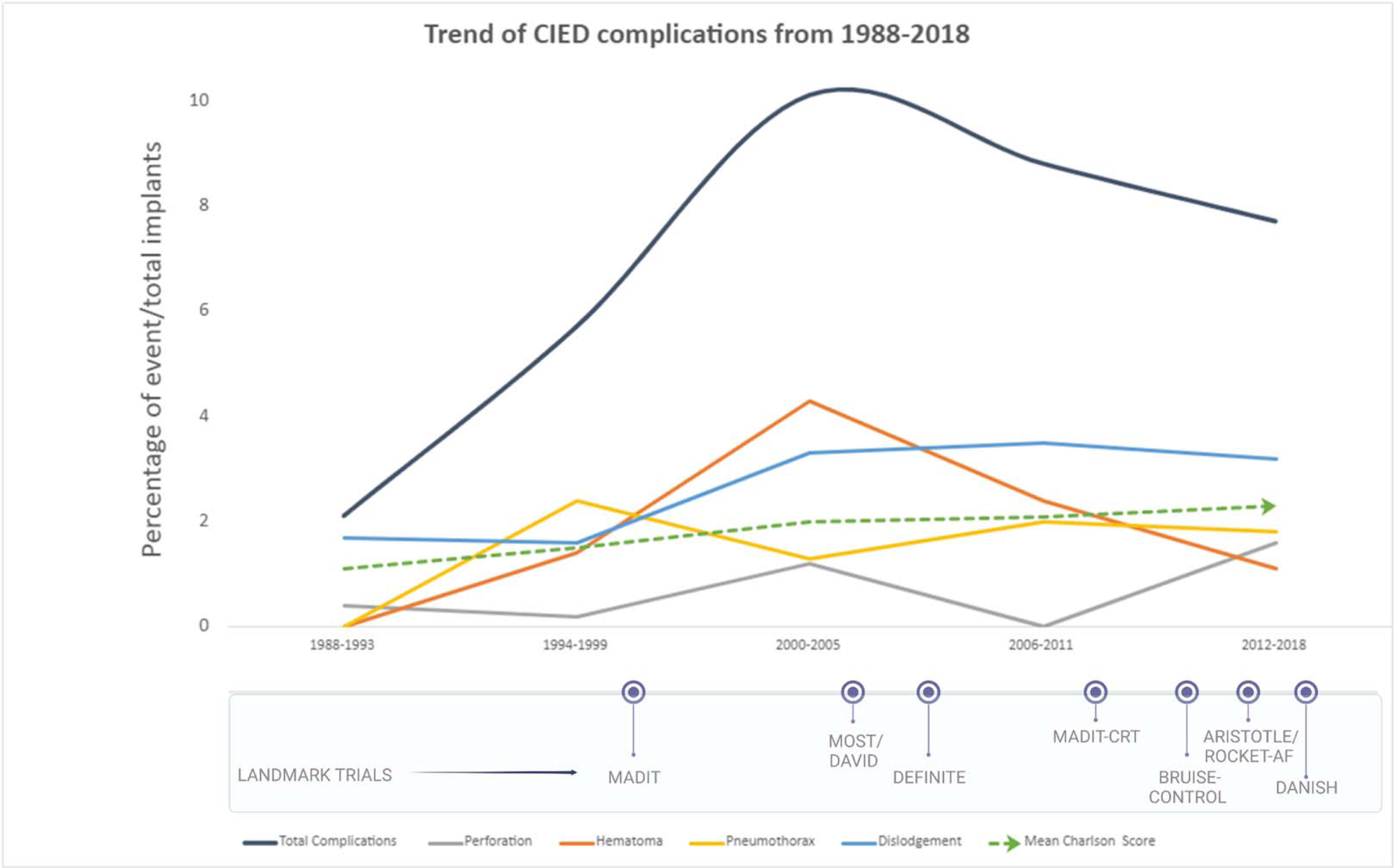
Central illustration. Trends of overall CIED complications from 1988-2018. Trendlines are provided for perforations, hematomas, pneumothoraces, lead dislodgements, and the mean Charlson Comorbidity Indices. CIED = cardiovascular implantable electronic device. MADIT(Multicenter Automatic Defibrillator Implantation Trial); MOST (Multi-arm Optimization of Stroke Thrombolysis); DAVID (Dual Chamber and VVI Implantable Defibrillator); DEFINITE (Defibrillators in Nonischemic Cardiomyopathy Treatment Evaluation); CRT (Cardiac Resynchronization Therapy); ARISTOTLE (Apixaban for Reduction in Stroke and Other Thromboembolic Events in Atrial Fibrillation), ROCKET-AF (Rivaroxaban Once Daily Oral Direct Factor Xa Inhibition Compared With Vitamin K Antagonism for Prevention of Stroke and Embolism Trial in Atrial Fibrillation); DANISH (Danish Study to Assess the Efficacy of ICDs in Patients With Non-Ischemic Systolic Heart Failure on Mortality

In relation to total implants, the percentage of pacemakers implanted has steadily declined over the years. The age and sex-adjusted incidence of PPM implantation increased between 1988-93 and 2000-05; then decreased in 2012-2018. The age and sex-adjusted incidence of ICD implantation increased between 1988-93 and 2000-05, then decreased in 2012-2018. In contrast, the age and sex-adjusted incidence of CRT implantation continued to increase significantly from 2000-05 to 2012-18(Figure 2A)^22^. The complications trends showed an initial increase in these groups followed by a decline in more recent years, statistically significant within each group, p< 0.05 (Figure 2B).

**Figure 2:**
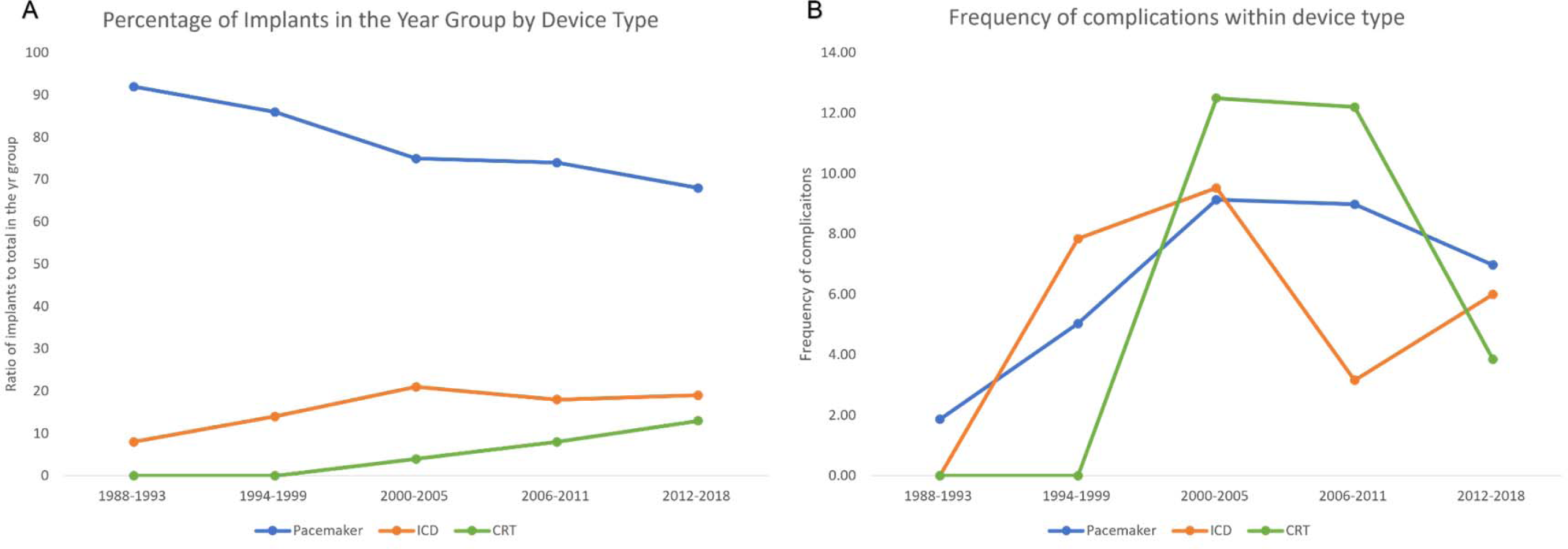
The trends of CIED complications by device type. (A) Percentage of CIED implants in the year-group by device type (B) Frequency of complications within device type. CIED = cardiovascular implantable electronic device; CRT = cardiac resynchronization therapy; ICD = implantable cardioverter-defibrillator; PPM = permanent pacemaker.

### Age and sex trends in CIED complications

When stratified by age, the percentage of implants for all periods was the lowest in patients < 50 years, higher in 50-70 years, and highest in patients > 70 years of age, p<0.01 (**Figure 3A**). The trend of complications in patients >50 years increased from 1988 to 2005 and then trended down (p<0.05). In patients under 50 years of age, the frequency of the complications rose from 1988-93 to 2000-2005, decreased in 2006-2011, and then increased again in 2012-18, although statistically not significant (**Figure 3B**).

**Figure 3:**
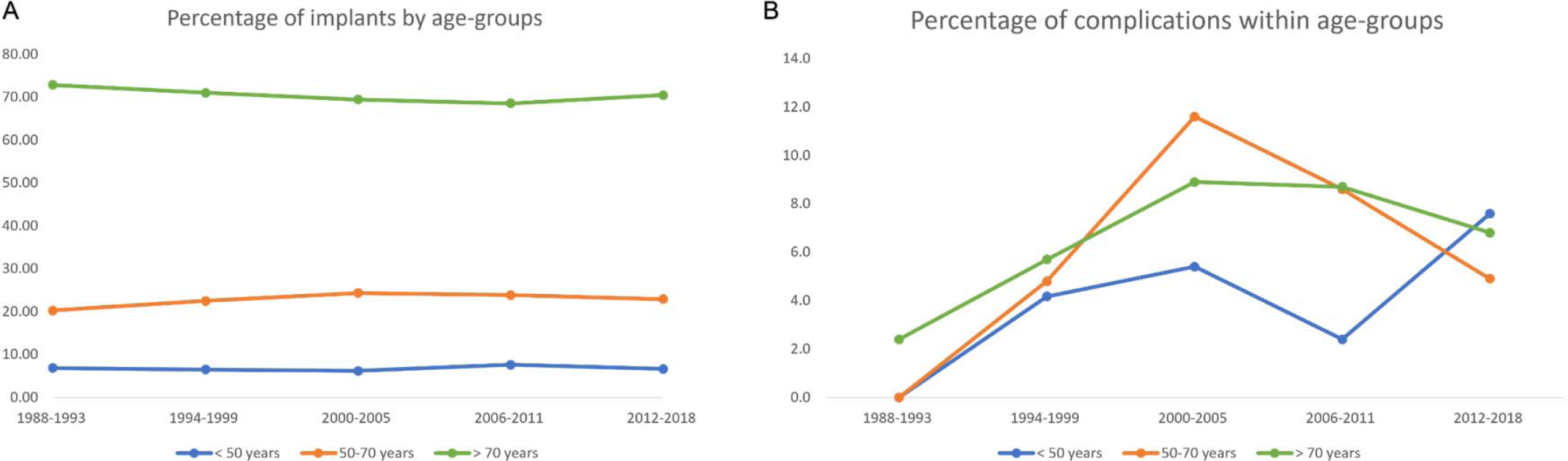
The trends of CIED complications by age. (A) Percentage of CIED implants in the year-group by age-groups (B) Frequency of complications within age groups. Abbreviations as in Figure 2.

Of the 175 patients with complications, 93 were males (53.1%), and 82 (46.9%) were females. Males have occupied a steadily growing percentage of the total implants in every time period. (**Figure 4A**). Within females, the complications increased from 1988-93 to 2000-05 and reached a plateau in recent years. In males, complications went up from 1988-93 to 2000-05 and then decreased until 2012-2018 (p<0.01) (**Figure 4B**).

**Figure 4:**
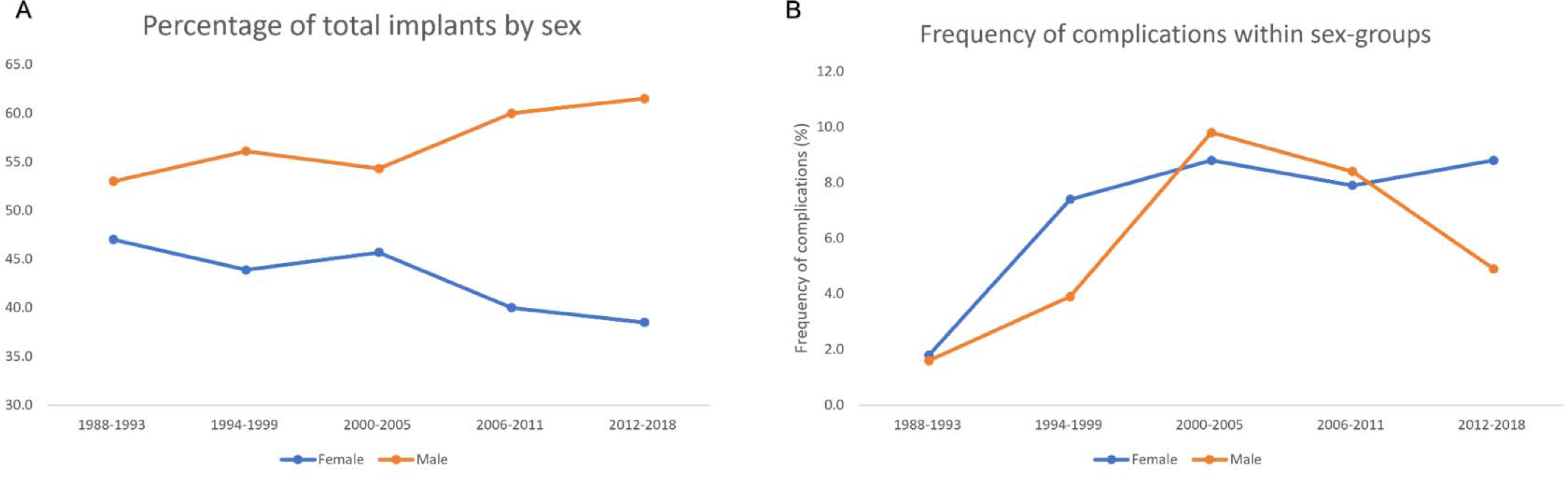
The trends of CIED complications by sex. (A) Percentage of CIED implants in the year-group by sex (B) Frequency of complications within sex groups. Abbreviations as in Figure 2.

### Trends in comorbidities and predictors of complications

Using the Charlson comorbidity index (CCI), patients’ comorbidities receiving CIED have increased over time (**Table 4**). However, there was no statistical difference between the CCI of patients with complications and those who did not within each time frame. When analyzed separately for each complication subtype, the only significant difference was noted in patients with hematoma from 2000-2011, who had higher CCI than those without. (**Supplemental Tables 2-5**). When controlled for other factors, utilizing a multivariate regression model, the presence of prosthetic valves, OR 1.62 (1.11, 2.37), p = 0.01, was independently associated with a higher risk of noninfectious complications. Obesity was independently associated with a lower risk of noninfectious complications, OR 0.68 (0.40, 0.95), p = 0.02.

**Table 4:** Charlson comorbidity index trends

| Year Group | Complications | Patients analyzed | Overall Mean Charlson Comorbidity Index * | Mean Charlson Comorbidity Index | P-value † |
| --- | --- | --- | --- | --- | --- |
| 1988-1993 | No | 225 | 1.1 ± 1.3 | 1.1 ± 1.3 | 0.20 |
|  | Yes | 4 |  | 0.3 ± 0.5 |  |
| 1994-1999 | No | 338 | 1.5 ± 1.5 | 1.5 ± 1.6 | 0.95 |
|  | Yes | 20 |  | 1.6 ± 1.1 |  |
| 2000-2005 | No | 525 | 2.0 ± 1.8 | 2.0 ± 1.8 | 0.51 |
|  | Yes | 53 |  | 2.2 ± 1.8 |  |
| 2006-2011 | No | 470 | 2.1 ± 1.8 | 2.1 ± 1.8 | 0.45 |
|  | Yes | 42 |  | 1.9 ± 2.1 |  |
| 2012-2018 | No | 729 | 2.3 ± 1.9 | 2.3 ± 1.9 | 0.90 |
|  | Yes | 47 |  | 2.3 ± 2.0 |  |

\* The p-value for the Charlson Comorbidity Index trend across all periods was significant at < 0.01
† The p-values described in this column are comparing patients with and without complication for each time period

### Overall survival after CIED implantation

The median follow-up was six years (interquartile range 3.0-10.9 years). Survival after CIED complication was 63.2% at five years (95% CI 55.9-70.4) and 35.4% (95% CI 27.4-43.4) at ten years, compared to 61.7% (95% CI 59.8-63.6), and 42.6% (95% CI 40.4-.44.8) respectively in patients without complications (p=0.07) (**Figure 5A**). After PPM, ICD, and CRT implantation complications, the survival was 35.8% (95% CI 26.7-44.9%), 38.2% (95% CI 18.9-57.5), and 15.6% (95% CI 0.0-43.2%), at ten years, respectively. In the PPM, ICD, and CRT implants without complications, the ten-year survival was 40.6% (95% CI 38.1-43.1, p =0.25) 53.0% (95% CI 46.5-58.55%, p=0.10), and 43.5% (95% CI 33.0-54.0%, p=0.67) (**Figure 5BCD**). Females and males with a complication did not significantly differ in survival compared to those without (p=0.21 and 0.20, respectively). (**Figure 6**)

**Figure 5:**
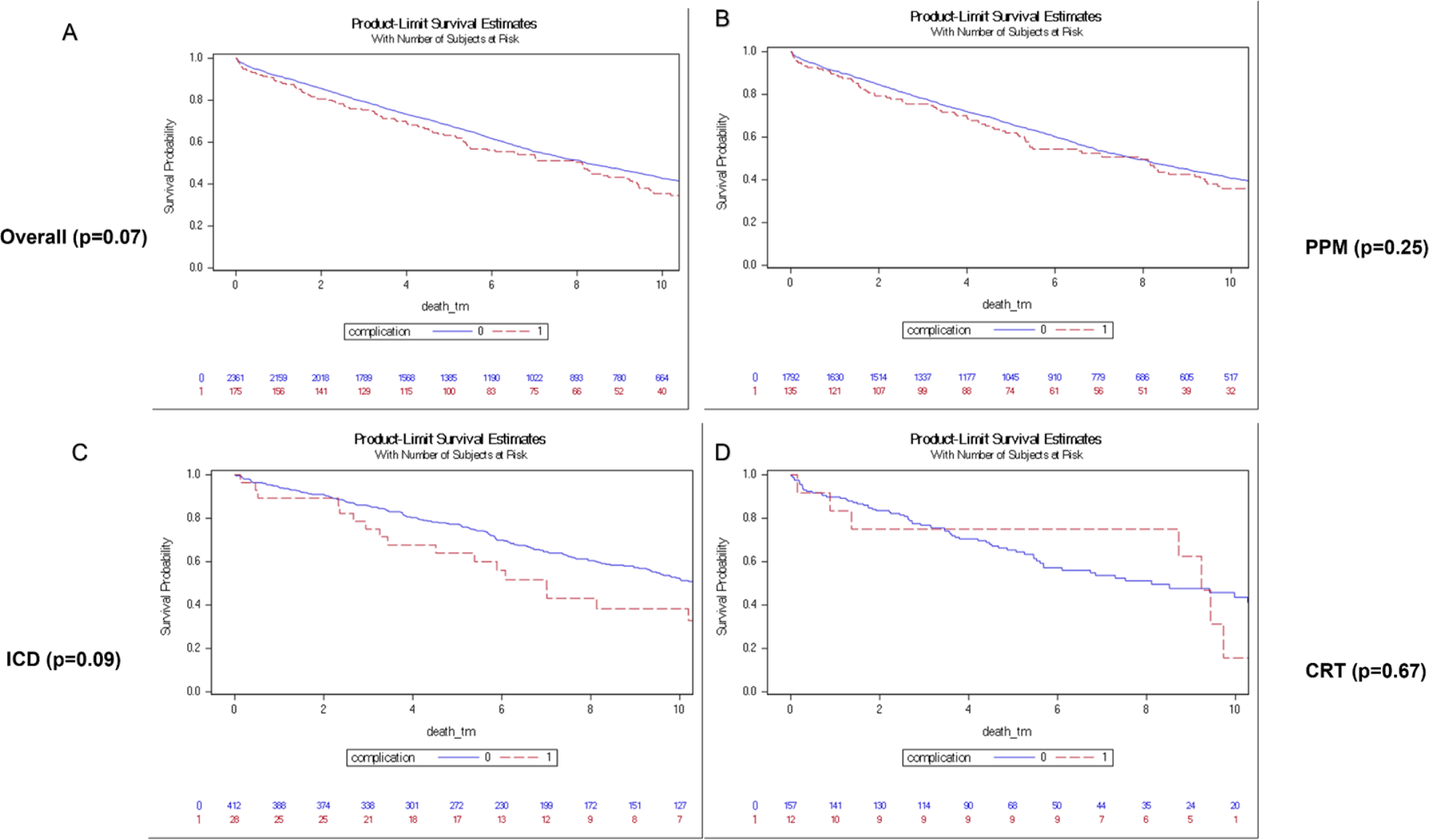
Survival Curves in (A) Total population, (B) PPM, (C) ICD, (D) CRT, based on the presence of complications or not. Abbreviations as in Figure 2.

**Figure 6:**
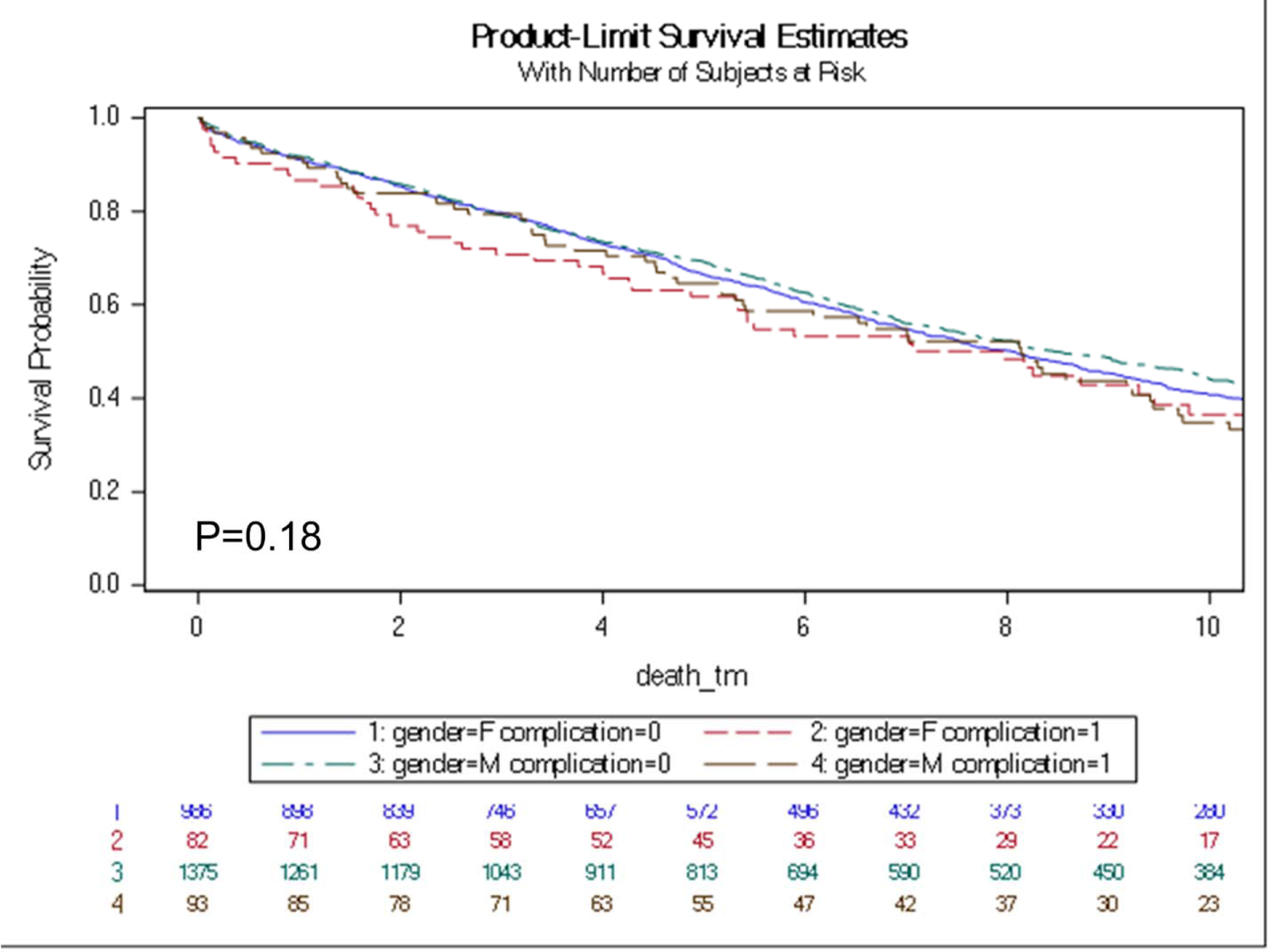
Survival curves by sex and complications. F = Female; M = Male.

**Table 5:** Multivariate regression model for predictors of complications

| Variable | Odds Ratio (95% Confidence Interval) | P-value |
| --- | --- | --- |
| Male sex | 0.82 (0.60, 1.14) | 0.24 |
| Age at event | 1.01 (0.99, 1.02) | 0.44 |
| Atrial fibrillation | 0.99 (0.68, 1.44) | 0.94 |
| Diabetes | 0.73 (0.53, 1.02) | 0.06 |
| COPD | 1.11 (0.80, 1.55) | 0.52 |
| CKD | 0.80 (0.57, 1.12) | 0.20 |
| Valvular Disease | 1.40 (0.91, 2.13) | 0.12 |
| Prosthetic Valves | 1.62 (1.11, 2.37) | **0.01** |
| Obesity | 0.68 (0.40, 0.95) | **0.02** |
| PPM implant | 0.91 (0.48, 1.70) | 0.76 |
| ICD implant | 0.93 (0.46, 1.90) | 0.85 |

Abbreviations – COPD: Chronic Obstructive Pulmonary Disease; CKD: Chronic Kidney Disease; PPM: Permanent Pacemaker; ICD: Implantable Cardioverter-Defibrillator

## Discussion

Using a large population-based record-linkage study, we describe trends in the noninfectious complications of CIED implantation. Our principal findings are (1) Lead dislodgement is the most common noninfectious complication after CIED implant, followed by hematoma, pneumothorax, and cardiac perforation. (2) The overall complications showed an up-trend from 1988 to 2005 and have reduced in frequency from 2005 to 2018. (3) The prevalence of complications is higher in patients over 50 years of age than younger patients. (4) The comorbidities of patients receiving CIED implants have steadily increased over time. (5) The presence of prosthetic heart valves was an independent predictor of noninfectious complications (6) Obesity tends to confer an independent risk-reduction concerning noninfectious CIED complications. (7) Long-term survival of patients with acute CIED complications was not different from those without complications but trended towards statistical significance.

### Incidence of noninfectious complications

The overall incidence of acute (30-day) noninfectious complications after CIED implants from 1988 to 2018 was estimated to be 6.9%, and that of major complications requiring intervention was 3.8%. Prior studies have shown wide variability in the percentage of complications (7-18%) due to heterogeneity in study populations, the definition of complications, data sources, and follow-up duration. ^10, 11, 19, 23–25^ Our findings seem to correlate with those reported by prior population-based studies ^23, 25^. However, there exists a discrepancy between the results of population-based studies, those obtained from administrative and claims databases ^11, 26, 27^, National ICD Registry ^18, 28^, and those from large-scale clinical trials. ^10, 17^

While clinical trials are gold-standard for assessing the effects of an intervention, the inclusion criteria are often quite strict, rendering a challenge in the generalization of observed events to the population. ^29^ Largescale databases, while reflecting the real-world scenario, are fraught with issues regarding the accuracy of data and lack of detailed follow-up. Therefore, a population-based medical record linkage system such as the REP serves as an intermediary and is crucial for mitigating the limitations of alternative data sources. The definitions used in our study for the complications were more inclusive than prior studies as our goal was to identify major and minor complications, and their predictors.

### Trends in device complications

Our group has previously described that the incidence of CIED implants in Olmsted County increased gradually from 1988 to 2005. However, from 2005 to 2018, a decline in CIED implants has been driven by a reduction in PPM and ICD implants^22^. The overall trend of CIED complications has been similar, increasing from 1988-93 to 2000-05, followed by a downtrend until 2012-18. The individual trends of lead dislodgement, hematoma, pneumothorax, and perforation followed the overall trends with slight variations when they peaked in the mid-2000s. While no prior studies have encompassed three decades of data, the trends seem to follow the most extensive reports from each period. ^10, 17, 18, 23, 25, 30^ These data appear closely related to the previously described trends with *infectious* CIED complications ^14^. The initial rise in complications seen from 1988 to 2005 reflects the increase in PPM in an aging population and expanding ICD indications for sudden death prevention, and the increase in the comorbidity burden^14, 22^. Improved implantation techniques, use of micropuncture needles, and imaging-guided access, with better periprocedural anticoagulation strategies, may have contributed to the subsequent decline in complications ^31–33^.

### Predictors of complications

Notably, the mean age of CIED implants at our institution was higher than in most prior population studies and trials^14^. Furthermore, our patients with 77% coronary artery disease, 75% valvular disease, 74% heart failure, 71% atrial fibrillation, and 62% diabetes mellitus, among others, represent a much sicker baseline population than prior studies^18–20, 23, 34^.

The presence of prosthetic heart valves was an independent predictor of noninfectious complications after adjusting for other implicated factors in device complications. These could be attributed to longer procedure times due to intricate and complex anatomy and the higher propensity to be on anticoagulation. The incidence of hematoma was reduced after 2011, likely reflecting the advantage of uninterrupted oral anticoagulation, a lower risk of pocket bleeding compared to heparin bridging. ^35^

Interestingly, obesity was shown to have an independent protective effect against noninfectious complications. The *reverse epidemiology* or *obesity paradox* is a well-established entity in chronic disease conditions and has recently been extrapolated to thoracic surgeries and CIED implants.^36–39^ Several mechanisms have been postulated to explain the obesity paradox, but the most relevant for CIED outcomes appear to be less cardiac cachexia, improved nutritional status, and better metabolic reserve with lower sarcopenia.^36, 39^ Obese patients tend to have a higher prevalence of comorbid conditions such as hypertension, diabetes, and sleep apnea that require closer periprocedural monitoring and may contribute to improved outcomes. This phenomenon is further evidenced by the extension of the obesity paradox to the critical care literature.^40^ It is important to note that the protective effects of obesity are likely lost at extreme body mass indices (>40 kg/m^2^), due to an increase in procedural time and complexity, with increased fluoroscopy duration.^23^

### Survival after device implantation and sex considerations

Data regarding long-term survival after CIED complications are sparse. We describe no significant difference in the 5-year and 10-year survival in patients without acute noninfectious CIED complications compared to patients with complications, although trending towards statistical significance.

Prior studies have shown that females are more likely to have short-term CIED complications.^23, 41^ While we saw a higher proportion of women in the patients that suffered a pneumothorax or perforation in our study, female sex was not an independent predictor of complications when adjusted for age and comorbid conditions. Additionally, females with acute CIED complications had no difference in long-term survival compared to males.

### Limitations

Despite the benefits of having access to extensive population-based linkage data, our study has several limitations. Akin to any observational study, we cannot rule out the confounding effect due to unmeasured variables. Our sample consists predominantly of Caucasians, especially among the elderly. Prior studies have suggested that the data obtained from the REP could be generalized to a large percentage of the United States population. This may be inconsistent, especially when applied to populations with a higher proportion of ethnic minorities. We were also limited in using ICD codes for CIED implantation, which may have underestimated the actual counts of procedures and comorbid conditions. Yet, the number of CIED implantations is consistent with previous REP studies with manual verification of all implantations and complications.

Although our data is population-based, most of the CIED-related care during the study period was provided by a single large tertiary hospital, which may have different practice patterns than community-based practices. While we understand that initial implants have a different risk profile than revisions, and generator replacements, we did not have a significant number of complications in lead revisions or device upgrades to make meaningful comparisons to de novo implants.

## Conclusions

CIED implant complications cause morbidity and may require re-intervention, but not associated with long term mortality. The frequency of acute noninfectious complications has been gradually decreasing in recent years, likely owing to safer practices. Prosthetic heart valves appear to be an independent risk factor for complications, while obesity seems protective, supplementing the growing obesity paradox literature. Close monitoring of the trends and consistent vigilance would be needed to maintain the downtrend of CIED complications.

## Data Availability

We will be happy to share any data or analysis as requested

## Abbreviations

CIED: Cardiac Implantable Electronic Devices
ICD: Implantable Cardioverter-Defibrillator
PPM: Permanent Pacemaker
CRT: Cardiac Resynchronization Therapy
CRT-P: Cardiac Resynchronization Therapy – Pacemaker
CRT-D: Cardiac Resynchronization Therapy – Defibrillator
FDA: Food and Drug Administration
REP: Rochester Epidemiology Project
ICD-9/10: International Classification of Diseases 9^th^ /10^th^ Revision
CPT: Current Procedural Terminology
NIS: National Inpatient Sample
NCDR: National Cardiovascular Data Registry

